# One does not fit all: Detecting work-related stress from mouse, keyboard, and cardiac data in the field

**DOI:** 10.1101/2025.08.02.25332538

**Authors:** Mara Naegelin, Raphael P. Weibel, Jasmine I. Kerr, Florian von Wangenheim, Victor R. Schinazi, Roberto La Marca, Christoph Hoelscher, Urs M. Nater, Andrea Ferrario

## Abstract

**Background:** Continuously and unobtrusively monitoring work-related stress may help combat its detrimental effects on mental and physical health. For work in offices specifically, mouse and keyboard data have been suggested as highly suitable data sources for stress detection, in addition to physiological data such as heart rate variability (HRV). However, previous studies have yielded mixed results regarding the potential of mouse and keyboard data to detect stress, and very few works have examined the connection in real work environments. Moreover, concerns regarding the robustness and validity of existing stress detection approaches have been raised, emphasising the need for more rigorous investigations in the field.

**Methods:** We conducted an 8-week observational field study with office employees (*N* = 36) where we collected mouse, keyboard, and cardiac data as well as self-reported stress during working hours. We derived mouse movement, keystroke dynamics, and HRV features, and trained regression models to detect self-reported stress levels using machine learning algorithms including Elastic Net, Random Forest, eXtreme Gradient Boosting (XGBoost), and Recurrent Neural Networks in two distinct modelling approaches: (1) one-fits-all, and (2) personalised. The first approach aims to detect the stress levels of a participant using training data from other participants. In the second, individual models are trained per participant.

*Results:* The one-fits-all modelling approach yields modest correlations with true labels (Spearman’s *ρ* = 0.078) under leave-one-subject-out cross-validation, with a slight improvement when incorporating time series of feature values (Spearman’s *ρ* = 0.096). In the personalised approach, XGBoost models trained on mouse and keyboard features reach an average Spearman’s *ρ* of 0.188 under blocked cross-validation. When optimised across machine learning models and feature sets, performance of the personalised approach further improves, reaching an average Spearman’s *ρ* of 0.296.

*Conclusion:* Our results suggest that that developing robust and valid stress detection models from in-field data remains challenging, reflecting the complexity of affective computing in naturalistic settings. Personalised modelling approaches show encouraging potential and warrant further exploration. We offer actionable recommendations to advance research on automated stress detection in real-world settings and openly share our dataset to promote innovation and collaboration within the research community.

## 1 Introduction

Work-related stress has become increasingly prevalent in the last decades [97, 1, 38]. The recent COVID-19 pandemic has aggravated the situation, with remote working blurring the lines between work and leisure time and increasing social isolation [120]. Prolonged or repeated experience of work-related stress can lead to a wide range of health-related issues, including burnout, depression, cardiovascular diseases, and metabolic disorders [22, 73, 53, 62]. Detecting and managing work-related stress early on is essential to prevent these physical and mental diseases and disorders and, thereby, reducing costs associated with presenteeism and absenteeism [38]. In particular, continuous stress detection can aid in self-monitoring, promoting self-awareness, and enabling individuals to manage stress effectively [71, 81].

Continuous stress detection is powered by machine-learning-based prediction models, which aim to detect stress levels from unobtrusively collected data [41]. A wide range of physiological or behavioural measures have been investigated as potential data sources [2, 43, 15, 108]. For office environments, where workers often spend the majority of their day interacting with their computer, researchers have suggested mouse and keyboard usage data as some of the most suitable data sources due to their unobtrusiveness, availability and cost-effectiveness [19, 25, 75]. Yet, despite such endorsements, relatively few studies exist investigating the use of mouse and keyboard data for stress detection. In fact, most of the current works have been exploratory in nature and conducted under laboratory conditions [25, 89, 84, 4, 35, 36]. Moreover, while some researchers have achieved promising detection results [89, 84, 4], others report only chance-level performance, casting doubts on the strength of the link between experiencing stress and mouse and keyboard usage behaviour [35, 36]. Cardiac activity data have also been proposed for stress detection specifically in work environments [75] since they can be collected with current consumer-grade wearables [46] and allow for the calculation of heart rate variability (HRV) measures that are widely recognised as established biomarkers of stress [9, 65]. In particular, they are among the most frequently used data sources in stress detection studies [46, 70]. Despite the potential shown by mouse, keyboard, and cardiac activity data, and although combining different data sources has shown the potential to improve the performance of stress detection models [2], literature exhibits a lack of field studies investigating the use of mouse or keyboard activity in combination with cardiac data to detect stress [82].

In general, given the considerable variability in terms of measurements, methods, and outcomes exhibited by stress detection studies, researchers have started to raise concerns regarding the robustness, validity, and generalisability of existing models, and emphasised the need for more rigorous investigations, especially in real-life contexts [80, 27, 124, 51]. One criticism pertains to the tendency of researchers to binarise labels into two classes (“stressed” versus “not stressed”), even though the intensity of stressors and the stress response can vary greatly [8, 80]. They argue, for example, that future research should avoid simplifying granular stress scores into two to three discrete classes, should cease to optimise accuracy at the expense of robustness and generalisability, and strive to report performance metrics alongside suitable baseline comparisons [12, 124]. Moreover, despite the high inter-individual variability in the stress response, most studies aim to develop a universal model that can detect the stress levels of all participants [99, 124]. Then, model performance is frequently assessed through sample-level cross-validation procedures which do not allow for an unbiased estimate of the performance on individual participants nor its generalisation to unseen participants, as opposed to, e.g., leave-one-subject-out cross-validation [12, 103]. Here, some researchers argue that a general, one-fits-all model for stress detection might never reach satisfactory results under real-world conditions, and advocate for more personalised approaches [117, 82, 128]. However, these have been applied less frequently in existing works, especially in real-world contexts [115, 23, 117]. Finally, deep learning methods such as recurrent neural networks (RNN), which take into account the inherent time series nature of stress detection data, also have been proposed to improve stress detection performance [12, 35, 107]. However, their application so far has been exploratory and limited by the availability of training data and thus warrants further investigation in real-world contexts. In summary, while still in its early stages, research on automated stress detection in real-world settings faces significant methodological challenges that have slowed its progress and constrained the performance of its modelling outcomes. These challenges are emphasised by the limited availability of publicly accessible datasets to investigate this topic. In fact, these datasets are essential for testing methodologies, validating previous findings, addressing modelling limitations, and fostering collaboration within the field. However, existing datasets, such as WESAD, SWELL-KW, and NEURO [102, 63, 10], are generated in controlled laboratory settings, while real-world data, such as those in [12], remain inaccessible to researchers.

In this work, we address these key gaps in research on detecting work-related stress using mouse, keyboard, and cardiac activity data in real-world settings. To do so, we present and discuss the results of an 8-week observational study conducted with office employees (*N* = 36) of a large insurance company. Self-assessed psychological states were captured several times daily per participant. We collected data on employees’ keyboard and mouse usage with a locally installed computer application, while employees’ cardiac activity was recorded with a wearable device. Then, we used machine learning (ML) modelling to examine to what degree the self-reported stress level can be predicted from the collected multimodal data. As a result, to the best of our knowledge, this work is the first to investigate the ML-based detection of work-related stress from mouse movement characteristics, keystroke dynamics, and HRV features in the field, marking a departure from traditional laboratory-based research. The contributions of this work to advancing ML-based stress detection in real-life contexts are as follows:

1. **We develop robust machine learning pipelines to detect work-related stress in the field** comprising (1) a one-fits-all, and (2) a personalised approach. We use different algorithms (Elastic Net, Random Forest, eXtreme Gradient Boosting, and Long Short-Term Memory network) and feature modality combinations, considering the self-reported stress level as continuous labels rather than discretising them into classes. We optimise and evaluate model performances using leave-one-subject-out cross-validation for the one-fits-all strategy and blocked cross-validation for the personalised one.
2. **We provide actionable guidelines for advancing automated stress detection in real-world settings**, tackling the numerous limitations that impede modelling performance in this field. In particular, we emphasise: (1) improving data collection and processing methods to ensure ecological validity in workplace environments, (2) designing robust machine learning pipelines that leverage both traditional and deep learning methods.
3. **We foster collaboration and innovation in technology-driven, in-field stress research, by making our dataset publicly available**, enabling other researchers to apply and test their methodologies. Data are added to a repository in the Open Science Framework (OSF) under https://osf.io/qpekf/ and are available upon request.

This study advances the stress detection domain by tackling the practical challenges of monitoring stress in real-world workplace settings using unobtrusive data sources, emphasising the superiority of personalised models over one-size-fits-all approaches. By offering actionable recommendations for future research and openly sharing its dataset, we aim to ensure a broader impact on the field, fostering innovation and collaboration toward the development of robust stress detection systems suitable for real-life deployment. These systems can help promote employee well-being, while mitigating the adverse effects of workplace stress on their mental and physical health.

## 2 Background

### 2.1 Stress and stress responses

Work-related stress refers to the psychological, physiological and behavioural strain that can arise in response to stimuli (or stressors) in one’s work environment [45], specifically in situations where the job demands are perceived to exceed one’s resources [66, 5]. Experiencing stress sets off a whole chain of cognitions, emotions, physiological reactions, and behavioural adaptations in order to enable us to cope with challenging circumstances [66, 24]. The field of stress research has developed tools and equipment to measure stress on these various levels [29]. The psychological response, for instance, can be captured with questionnaires granting insights into the various cognitive and emotional processes reflective of stress [32].

On a physiological level, blood and saliva samples provide information on the activation of both the autonomic nervous system (ANS) and the hypothalamic-pituitary-adrenal (HPA) axis by determining the levels of cortisol, epinephrine, norepinephrine or salivary alpha-amylase [57, 29, 3]. Alternatively, other physiological markers may be used to assess stress reactivity in a less obtrusive manner. These biomarkers—e.g., quantifying cardiac, electrodermal or muscle activity— typically reflect the stress reactivity of the sympathetic and/or parasympathetic nervous systems (SNS and PNS), the two branches of the ANS [9, 26, 116]. Here, heart rate variability, especially, has been established as an important indicator of stress, health, and disease [50, 77, 61]. HRV refers to the variation in time between successive heartbeats, also known as the R-R intervals [9]. Wearable devices that can monitor these physiological markers have become increasingly affordable, comfortable, and accurate in recent years [43].

Behavioural indicators of the stress response have been researched less thoroughly, but have started to gain popularity because they can often be collected completely unobtrusively and, in some cases, without the need for expensive additional equipment [2]. For example, stress may lead to measurable changes in facial expressions and body posture, but also computer interactions and mobile phone usage [2, 19]. In contrast to the psychophysiology of stress, the majority of existing works on the effects of stress on technology usage have been exploratory and a-theoretic in nature. Here, Neuromotor Noise Theory [122] and Attentional Control Theory [34] have been proposed as a basis for understanding the relationship between these data sources and stress [35, 36, 6, 84]. Van Gemmert and van Galen’s Neuromotor Noise Theory suggests that stress increases the degree of neuromotor “noise”, the variability in neural and physiological signals, which leads to somewhat imprecise motor control and movements [122]. This theory supports the use of behavioural data generated through interactions with mouse and keyboard devices in various studies aimed at automatically detecting affective states, including stress, anxiety, and emotions such as confidence and nervousness. These investigations, conducted primarily in laboratory settings, demonstrate the potential of such interactions as reliable indicators of emotional and psychological state [129, 100, 123, 33]. In particular, Neuromotor Noise Theory has been used to show that stress is associated with a speed-accuracy trade-off in computer mouse movements [6]. Similarly, Attentional Control Theory proposes that anxiety (also an affective response to stress) presents itself in impaired attentional control affecting motor function [34]. Consequently, stress researchers have turned to readily available data sources at the workplace such as mouse movements and keyboard dynamics believed to capture stress-related changes in motor noise and attentional control [19]. Furthermore, the non-invasiveness and widespread availability of devices like mouse and keyboard make them more affordable and less intrusive sources of behavioural data in the workplace compared to physiological sensors or cameras [19]. Mouse and keyboard devices are characterised by high levels of cost-effectiveness, transparency and user privacy, and feature diversity [19]. Lastly, contextual data such as calendar entries, ambient sound, location, or even weather information have also been used to detect stress [2, 19]. It is worth noting, however, that the context does not change based on a person’s stress levels, rather, it may correlate with or itself contribute to a higher or lower probability of stress directly or indirectly [2].

### 2.2 Stress detection studies: From lab to field

Initially, stress detection research focused on laboratory studies to explore and test different data modalities and modelling approaches under controlled conditions, where stress can be deliberately elicited and potential confounding factors are easily eliminated or at least minimised [15, 124, 84, 85]. Many of these laboratory studies have yielded promising results (for reviews, see, e.g., [2, 43, 15, 41]). In recent years, a growing number of field studies attempted to replicate the promising findings from laboratory experiments in real-world settings. However, the complexity of real-world environments poses significant challenges. As a result, stress detection field studies have generally achieved substantially lower performance levels than their laboratory counterparts [111, 15]. In the following, we outline some of the main challenges faced by stress detection field studies and discuss the relevant related works and their limitations.

In ecological settings, stress is not elicited with artificial, standardised stressors, such as the Stroop test or mental arithmetic tasks, but rather occurs naturally and unpredictably [15]. Stress responses may thus also vary considerably in both intensity and manifestation, since different stressors may differentially stimulate various aspects of the stress response [110, 113]. Moreover, the “ground truth” of a person’s stress level cannot be as easily established as in laboratory experiments, where the presence or absence of a stressor is determined by the experiment protocol. In field studies, researchers thus rely almost universally on Ecological Momentary Assessments (EMA) for capturing subjective self-reports of stress [111, 27]. However, EMAs can only be sampled at discrete time points and require active participation from the study subjects, so they might fail to capture a significant portion of stress events [27] and impose limits on the amount of labelled data that can be collected with reasonable effort [118]. In particular, well-validated but time-consuming multi-item instruments to assess perceived stress are often forgone in favour of simpler and shorter scales [101, 115, 121], which might risk lower reliability and higher inter-subject variability [114].

Data in field studies are much more prone to noise and missing data, and suffer from an abundance of potential confounds [111]. This complicates the data processing steps such as feature extraction, and necessitates the adequate handling of missing data. In addition, the predictive potential of a data source is not the only aspect which needs to be taken into consideration in real-world contexts [19, 84]. Researchers must also weigh factors such as the degree of privacy invasion and obtrusiveness a data source poses, in addition to the cost and scalability of the requisite hardware or software [19]. Here, data sources such as mouse, keyboard and cardiac activity have been rated by employees as more acceptable than others, e.g., video data used to estimate facial expressions or body postures, for the purpose of real-time stress detection [59, 56].

## 3 Related work

In order to detect stress automatically and in real-time, researchers apply ML algorithms to one or more data sources [41]. In the following, we outline the related works on stress detection using cardiac, mouse and keyboard data, and the challenges and gaps in existing ML modelling approaches.

### 3.1 Stress detection from cardiac data

Cardiac activity has arguably become one of the most used data sources to detect stress [111] for two reasons. First, HRV has long been established as an important indicator of stress, health, and disease [50, 77, 61] and has been used successfully to distinguish between stressed and relaxed states in lab studies (e.g., [49, 90]). Second, wearable devices equipped with photoplethysmography (PPG) sensors, such as consumer-grade fitness trackers and smartwatches, are becoming increasingly accurate, affordable, and comfortable [43, 46]. Nevertheless, a recent study by Martinez et al. [74] found the link between HRV and perceived stress in the field to be greatly diminished. In fact, HRV measures explained only 2.2% of the variance in self-reported stress in their field study monitoring 657 participants over eight weeks. Similarly, Smets et al. [112] found the performance of their models to be insufficient for practical application in a large-scale ambulatory stress detection study utilizing HRV and other wearable-based physiological data, while Booth et al. [12] argue that weak associations between behavioural signals and subjective mental states are to be expected in real-life contexts. More research is thus needed to clarify the potential of HRV data from consumer-grade equipment for stress detection in the field [75, 54].

### 3.2 Stress detection from mouse and keyboard data

On the behavioural side, technology interaction data, e.g., from smartphones or computers, have been proposed as some of the most suitable options for ML-based stress detection in the field, since they can be collected without any expensive or obtrusive additional equipment [19, 2]. While these data only arise during users’ interactions with the device, knowledge workers generally spend the majority of their working hours in front of a computer [60], making computer interaction data such as mouse and keyboard activity uniquely suited for stress detection in office environments [19, 25, 75]. Despite this, research investigating mouse and keyboard data for stress detection is still relatively scarce and consists almost entirely of lab studies [56]. Further, the empirical evidence from lab studies ranges from promising to inconclusive. For example, Pepa et al. [89] could detect three stress-level classes from keyboard (76% accuracy) and mouse (63% accuracy) data from 67 participants who completed a series of computer tasks while exposed to stress-eliciting sounds and other disturbances. In contrast, Freihaut and Göritz found that self-reported stress was neither consistently linked with keyboard data [36] nor with mouse data [35] in their recent laboratory experiments. In the first study to investigate the link between mouse data and work-related stress in the field, Banholzer et al. [6] collected mouse movement data of 70 employees for 7 weeks and found a significant two-way interaction effect of mouse movement speed and accuracy on self-reported stress.

### 3.3 Multimodal stress detection

While researchers have found that the combination of different modalities generally improves the performance of stress detection models [2, 64], very few studies combine mouse and keyboard data with physiological data sources and, in particular, cardiac data. Naegelin et al. [84] simulated a group office environment and an experimental protocol involving a baseline workload and work-related stressors. On the data from 90 participants, they achieved an F1-score of 0.63 when predicting low, medium and high self-reported stress based on mouse and keyboard data. Interestingly, they found that adding HRV data to the models did not improve performance. Two studies have used a small number of mouse and keyboard features (e.g., the number of mouse button clicks and number of pressed keys) in combination with a whole range of features from behavioural, physiological and contextual data sources, making it difficult to draw any isolated conclusions regarding the potential of mouse and keyboard modalities. Namely, Sanchez et al. [101] monitored 57 participants during 5 working days, collecting heart rate, computer interactions, physical activity, and sleep data. They achieved 79% accuracy when detecting low, medium and high self-reported stress, but since the train-test split was performed after oversampling the minority classes on the whole data set, this estimate might be somewhat optimistic. In the study by Morshed et al. [82], 46 employees provided email, calendar, computer interaction, and webcam data, as well as regular self-reports on their sleeping, eating and drinking habits during four weeks. The camera data was used to derive facial expressions and HRV features. When predicting low versus high stress based on these combined data sources, the authors reached an average F1-score of 78%. Furthermore, participants rated mouse and keyboard data as the most acceptable of collected sources.

### 3.4 Robustness and validity of stress detection models

Stress detection research has yielded inconsistent results across studies, with large discrepancies in study design, collected measures, employed methods and approaches contributing to a lack of consensus in the field [12, 87, 15]. This diversity has led some researchers to call into question the validity and generalisability of existing stress detection models [35, 12, 27, 124, 51, 84]. Indeed, in their systematic literature review on stress detection from wearable devices, Vos et al. [124] found that the majority of surveyed studies were of low to medium quality, had questionable statistical power and did not sufficiently address model generalisation. In a recent commentary, D’Mello and Booth [27] discuss the sobering results from the Multimodal Objective Sensing to Assess Individuals with Context (MOSAIC) program initiated by the U.S. government, which found the performance of automated methods to detect self-reported daily stress from physiological, behavioural, and context data to be near-zero under rigorous, independent testing. In an effort to bridge the discrepancy between previous high-performing approaches and their own findings, the group around D’Mello and Booth have developed guidelines for more robust and valid stress detection models [12]. In line with [75, 124, 80], they argue against the oversimplification of stress labels into binary classes and further emphasise that any quantisation of stress level scores may lead to a loss of relevant information and diminish construct validity [12]. In addition, previous works have been criticised for liberally excluding lower-quality data or discarding mid-range stress labels in an effort to simplify the prediction problem, sacrificing robustness for better performance [12]. Finally, a lack of suitable baseline comparisons can make it difficult to properly assess achieved performance, especially since classes are often severely imbalanced [12, 40]. However, very few studies have investigated whether the differences in achieved model performances (between different approaches or to a selected baseline) were statistically significant [124, 12]. The models Booth et al. [12] developed following their framework based on collected physical activity, workplace behaviour, phone usage, sleep, heart rate, weather, and weekday data achieved small-to-medium correlations with daily self-reported stress when evaluated on unseen subjects.

### 3.5 Deep learning-based stress detection

Another point of contention is the potential of deep learning methods to improve on existing stress detection results. While many studies have employed more traditional ML methods such as k-Nearest Neighbours, Support Vector Machines, Random Forests or Boosting algorithms [41, 84], researchers have only recently begun to explore more complex methods. Here, one approach aims to leverage the temporal information available in sequences of collected data with the help of time-aware methods such as RNN. While some studies could demonstrate promising results, e.g., for physiological data-based stress detection [121, 107], others were less successful. For example, Freihaut and Göritz [35] found their CNN and RNN approaches on mouse movement data to perform on par with random guessing, while the RNN methods tested by Booth et al. [12] also yielded chance-level performance. Likely, the limited amount of available data across studies means that the potential of data-intensive deep learning methods has not yet been fully leveraged.

### 3.6 Personalised stress detection approaches

Regarding the generalisability to new users, other researchers go so far as to question the feasibility of a general one-fits-all approach for stress detection under real-world conditions [117, 82]. They argue that such an approach cannot sufficiently account for the unique characteristics and patterns of individual stress experiences and suggest personalised modelling approaches instead. For example, in the previously mentioned study by Morshed et al. [82], 78% accuracy was achieved for participant-dependent models, while a leave-one-subject-out cross-validation procedure yielded the much lower accuracy of 46%. Soto et al. [115] collected cardiac, electrodermal and physical activity, skin temperature, and respiration data from 14 participants over 8 weeks. They found that a general model performed worse than random guessing of low versus high stress states, while per-participant models could on average achieve a small improvement in accuracy over a stratified random model. The main drawback of personalised approaches, e.g., training an individual model per participant, is the need to collect training data for each new user [16, 111].

In conclusion, despite the substantial body of existing research on automated stress detection, open questions remain regarding the ecological validity, generalisability and robustness of modelling procedures, especially in field settings. More research is needed on the use of mouse, keyboard and cardiac data sources for stress detection in real-life work environments, as well as the implementation of comprehensive, robust ML modelling approaches, e.g., one-fits-all versus personalised, and time-aware methods such as RNN. We address these challenges in the field study presented in the forthcoming sections.

## 4 Methods

In this section, we describe the field study procedure and data collection protocol, followed by a description of the data processing and modelling steps and finally the evaluation of the developed ML models. We visualise our methodology in Figure 1.

**Figure 1:**
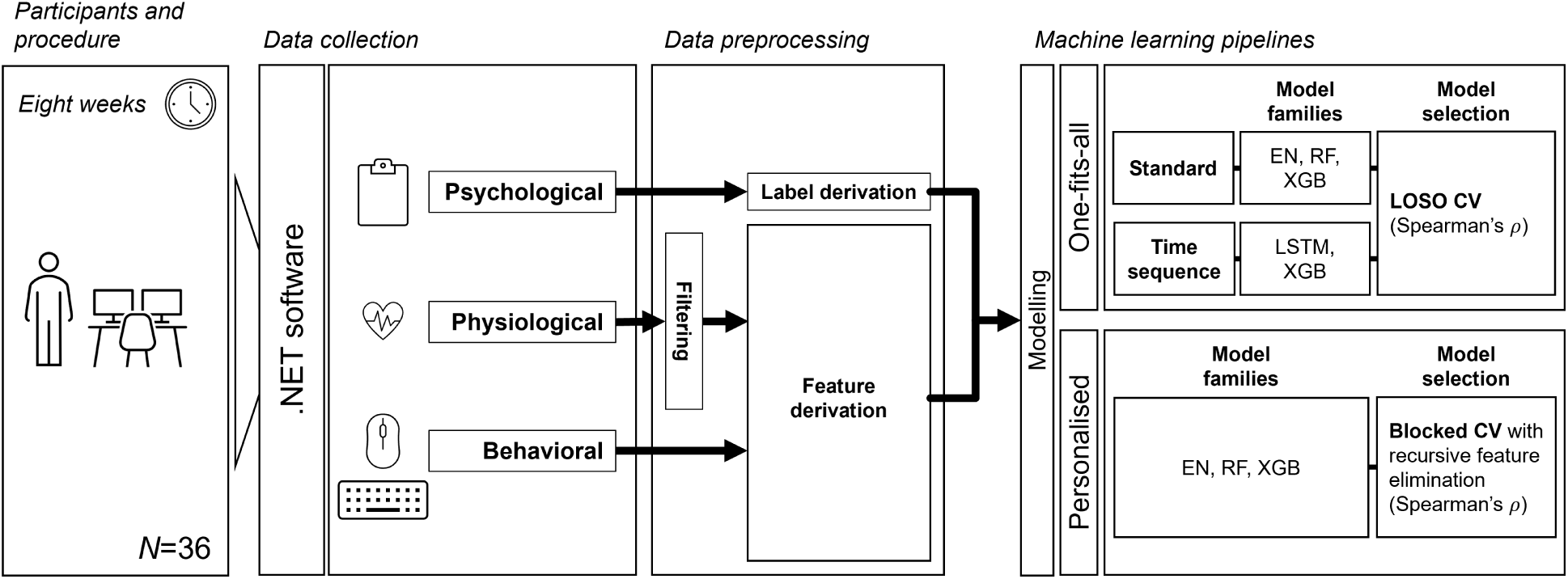
Our methodology for the automated detection of stress in a real-world office. We describe the step “Participants and procedure” in Section 4.1, “Data collection” in Section 4.2, “Data preprocessing” in Section 4.3, while the two approaches under “Machine learning pipeline” are introduced in Section 4.4. Abbreviations: EN = elastic net; RF = random forest; XGB = extreme gradient boosting; LOSO = leave-one-subject-out; CV = cross-validation.

### 4.1 Participants and procedure

Participants were recruited from the employee pool of a large insurance company (*>* 5000 employees) through a post on the company’s intranet. Interested employees were first informed of the content and requirements of the study and then asked to complete an online screening questionnaire to assess their eligibility for participation. Specifically, participants were required to be at least 18 years of age, be fluent in German, have normal or corrected-to-normal vision, use a computer for the majority of their working time, work for at least three days per week, and have no more than two consecutive weeks of planned absence during the data collection period. Furthermore, participation was not allowed in case of impaired use of arms or hands, intake of cardioactive medications (e.g., antidepressants, antipsychotics, anti-hypertensives), or consumption of psychoactive substances (e.g., LSD, Ecstasy, marijuana). The consumption of caffeine, alcohol or nicotine was not restricted.

In total, 42 participants meeting the inclusion criteria signed up for the study. Of these, three participants revoked their participation within the first week of data collection, and another three participants dropped out later during the data collection period due to a change in employment or personal reasons. These participants were excluded from the analysis, resulting in a data set comprising data from 36 different employees.

Participants who met the inclusion criteria were invited to individual onboarding meetings. There, an experimenter explained the procedure of the study in detail and answered potential questions. After participants gave their informed consent, they were guided through the installation and activation of the study hard- and software on their working laptops and instructed regarding their use during the study. The data collection for each participant started on the Monday following their respective onboarding meeting. We aimed for eight consecutive weeks of collected data. If a participant was absent from work for an entire working week or more (e.g., due to vacation, sick leave, etc.), we extended their data collection period by the respective amount of time—subject to the participant’s agreement and availability. The study took place from January to May 2022.

After the data collection period was completed, we held individual debriefing meetings with all participants, where we guided them through the de-installation of the data collection app from their local machine, instructed them to return the provided heart rate sensor and answered any further questions that might have arisen. Participation was not remunerated. Participants received a personal report on their own collected data, giving an overview of their self-assessed mental health, computer usage, and cardiac activity over the data collection period. The study was approved by the University’s Ethics Commission (EK 2021-N-209) and conducted in accordance with the Declaration of Helsinki.

### 4.2 Data collection

A self-developed .NET software was used to collect participants’ daily activity data. Specifically, the data collection app recorded participants’ mouse and keyboard activity, cardiac activity (via an optical heart rate sensor), and self-assessments of their current mental state. All collected data was temporarily stored on the participants’ local machine and regularly transferred directly to the university’s database server. Once the data was successfully transferred, it was removed from the local machine.

The cursor position during mouse movements (i.e., no position was logged if the mouse position remained the same), mouse button activity and keyboard activity were recorded in real-time. However, to adhere to the insurance company’s privacy and data protection regulations, the collected keyboard data was partially aggregated in-memory before being stored on the local machine (see Section 4.3.2 for more details).

Cardiac activity was collected via the optical heart rate sensor Polar Verity Sense (Polar Electro Oy, Kempele, Finland), which uses photoplethysmography to capture the time intervals between heartbeats (in ms, recording frequency of 135 Hz). The device is lightweight with an elastic strap and can be worn either on the lower or upper arm, depending on user preference. During the onboarding meeting, participants were instructed on how to wear and handle the device. Specifically, they were asked to turn on the sensor at the start of every working day and wear it whenever they were working on their computer. The battery of the Polar Verity Sense lasts up to 30 hours, so they were asked to charge their device every other night via a USB port (e.g., by plugging it into their laptop). The raw cardiac data collected by the Polar Verity Sense was continuously streamed to the data collection app on the participants’ local machine via Bluetooth, implemented using the Polar software development kit.

Participants were prompted to assess their psychological state several times a day via a small pop-up window on the lower right side of their screen. The pop-ups appeared semi-randomly, roughly 1 hour after they first started the data collection app, then 2, 3, and 2 hours after each previous EMA was provided. If they did not want to answer it, then they could close the pop-up window, and it would reappear 20 minutes later. They could also manually open and submit an EMA by clicking on the data collection app icon whenever they wanted. Each EMA consisted of Visual Analogue Scales with values from 0 to 100, similar to [121, 68, 67]. Specifically, participants were asked to rate their mental state in the previous hour along five dimensions: stress, valence, arousal, wakefulness, and exhaustion. For the purpose of this manuscript, we focus on the “stress” scale. High levels indicate a higher level of perceived stress. The EMA questionnaire used in this study can be shared upon request.

Finally, participants received weekly emails with a link to a questionnaire hosted on University servers. Here, they provided their work location (i.e., office or home office) for each half-day and filled in the Stress Scale of the Depression Anxiety and Stress Scales (DASS) [72] for the previous work week. The very first questionnaire also included sociodemographic questions (e.g., gender, position within the company, percentage of employment) and the Screening Scale of the short version of the Trier Inventory for the Assessment of Chronic Stress (TICS-2-K) [104], while the last questionnaire also contained the TICS, as well as an assessment of the user experience of the data collection system via the System Usability Scale (SUS) [13] and self-developed items regarding the obtrusiveness of individual components.

### 4.3 Data preprocessing

#### 4.3.1 Mouse data

The raw mouse activity data collected consists of a timestamp, the coordinates of the cursor on the screen (i.e., *x*- and *y*-position in pixels), and the type of mouse operation recorded. These operations are categorized into movements, clicks, or scrolling events, as defined in previous studies. Specifically, a movement is recorded anytime the cursor is displaced, i.e., if its coordinates have changed since the previous sampling of its location. Click operations are distinguished by the mouse button which was pressed (i.e., a left or a right mouse button click), and scrolling operations are characterised both by the direction (up or down) in which the scrolling wheel is turned.

All mouse features used in this study are grounded in relevant literature on stress detection, ensuring alignment with established methodologies [64, 129, 100, 84]. These features typically relate either to the occurrence of specific mouse events (e.g., number of left clicks or scrolling events) or characterise the dynamics of the observed mouse movements [129, 18, 4]. Examples of such mouse movement features are the covered distance, the cursor speed, the deviation from an optimal straight line between its start and end point, or the traversed angle. In particular, Naegelin et al. showed that the mean Euclidean distance between the end and start locations of mouse movements, and the count and standard deviation of durations in mouse movement pauses correlate with various stress-related affective states [84]. Further, Freihaut and Göritz’s review of the theoretical foundations and empirical evidence linking stress to mouse movement behaviour strongly supports the idea that stress affects goal-directed actions, including mouse movements [35]. While there is no clear consensus on the definition of a mouse movement [6, 100], following relevant literature, we define it as a consecutive series of mouse locations on the screen that is delimited by either a click or scroll operation or a pause of more than 1s, similar to [100, 84]. To derive features for a given time window of raw data, we segmented the mouse activity data into individual mouse movements, calculated a set of characteristics for each movement, and aggregated across the time window using the mean and standard deviation, as in [84, 129]. In total, we derived 53 mouse features, see Appendix B for the full list and description of these features.

#### 4.3.2 Keyboard data

Raw keyboard activity data consisted of a timestamp, the type of event (i.e., key pressed or released), and a key identifier. However, to comply with the privacy and security requirements of the participants’ employer, we programmed the data collection app to partially aggregate the raw keyboard activity data in-memory on participants’ local machines before transfer to the University’s database server. Specifically, three different kinds of partial aggregations were performed. They were chosen to prohibit any meaningful inference on the content of what participants have typed at any point during the study, while still allowing the derivation of common keystroke dynamics features for the modelling. First, the press and release events for each keystroke were logged with their respective timestamps rounded to seconds and replaced the key identifier with a random key ID. Second, the occurrence counts for specific relevant keys and groups of keys were recorded at 5-minute intervals. And third, we logged keystroke dynamics for three of the most common digraphs (‘en’, ‘er’, ‘ch’) and trigraphs (‘ein’, ‘ich’, ‘nde’) of the German language. Specifically, for each graph, we recorded the type of di- or trigraph and the time differences (in ms) between all press and release events of in the graph. Similarly to the key counts’ data, the graph data was recorded with a time granularity of 5 minutes. Due to a recording error, the second digraph keys were not recorded properly, limiting the available digraph features to the durations of the first digraph keys.

We derived a variety of keyboard features from the three types of partial aggregations of the raw data, explicitly following established methodologies from prior literature. These features have been previously validated and utilised in studies such as [100, 84, 64, 123, 33], ensuring that our approach aligns with and builds upon existing research in the field. For example, we derived key counts and various keystroke dynamics, which have been identified in previous studies as relevant indicators for stress detection [123, 33, 84]. Naegelin et al. recently demonstrated that these features, in particular the number of typing pauses, as well as their mean duration and standard deviation, are significantly correlated with various stress-related affective states [84]. First, we considered the occurrence counts for the different keys or key groups such as the number of letter keys or error keys pressed in a specific time window (of at least 5 min length given the aggregation level of the raw data). Second, we used the key presses per second to infer typing speed and typing pause characteristics (e.g., number and mean length of pauses). Here, a typing pause is a time interval of *>* 1s with no recorded keystrokes, based on previous work [125, 84]. And third, from the graph data, we derived key latency and flight times within each graph, aggregating across graphs of the same type by taking the mean and standard deviation [100, 84]. In total, we derived 49 keyboard features, see Appendix B for the full list and description of these features.

#### 4.3.3 Cardiac data

The raw cardiac data provided by the sensor consists of arrays of inter-beat-intervals (in ms), which are streamed to the data collection app every five seconds and written to the database with the timestamp of when they were received. Concatenating the arrays of inter-beat-intervals recorded during a given time window thus yields a Peak-to-Peak Interval (PPI) time series from which different HRV indicators can be derived. We used the Python package hrv (version 0.2.10, [7]) to preprocess the cardiac data. PPI data is often subject to considerable noise, for example, due to motion artefacts or ectopic heartbeats, and thus needs to be filtered before feature derivation [88]. Specifically, for each time window, we removed PPIs with extreme values (i.e., under 300 or over 2000 ms) and applied the threshold filter of the hrv package to detect outliers. This filtering method is based on the threshold algorithm of the popular HRV analysis software Kubios, which compares each inter-beat interval to the local median and excludes it if it differs by more than a certain threshold. We chose the threshold “medium” (250 ms) in our case as PPG data is fairly sensitive to noise [78]. Excluded values were replaced by cubic spline interpolations. However, if the filtering resulted in less than 50% of valid data remaining, the entire time window was seen as invalid and discarded [8]. From the denoised data, we calculated a range of time- and frequency-domain, as well as non-linear measures of HRV that are commonly used in literature [106, 90]. The frequency domain measures were derived from the Power Spectral Density (PSD) estimate of the detrended and interpolated (cubic spline interpolation at 4 Hz) PPI time series. The PSD was estimated via Fast Fourier Transform using Welch’s method with Hann windows. In total, we derived 9 HRV features, see Appendix B for the full list and description of features.

### 4.4 Machine learning pipelines

We derived the mouse, keyboard, and cardiac features as described in Sections 4.3.1 to 4.3.3 above on the 60-minute time windows preceding an EMA answer [109], and assigned the corresponding self-reported stress levels (ranging from 0 to 100) to each observation. Moreover, we included work location (home office, office), gender (male, female), and position within the company (employee, management) as binary input features. As a result of the data preprocessing, we obtained a total of 3574 labelled observations across 112 features (53 mouse, 49 keyboard, 9 cardiac, and 3 context features).

We investigated the performance of different regression models on either the behavioural features (mouse and keyboard; MK models), the physiological features (HRV; H models), or all three modalities (mouse, keyboard and HRV; MKH models). Some of the observations had missing values. We removed only observations with no predictive content, i.e., observations with only partially missing values were handled with data imputation techniques, as described further below. Specifically, for each feature subset, we excluded observations that contained no valid data from any of the included modalities (i.e., holding no information at all). This affected mostly the H models due to the data quality issues of the PPG data. Moreover, four participants had no valid PPG data at all throughout the entire data collection, and thus their data were excluded for the models which included the HRV modality in their feature subset (i.e., MKH and H models were trained and evaluated on data from 32 participants).

We used the Python packages scikit-learn, XGBoost and the Keras API of TensorFlow (versions 1.3.0, 1.6.1 and 2.12.0, respectively) to implement all ML models.

#### 4.4.1 One-fits-all approach

In the one-fits-all approach, models are trained with the aim of predicting the stress levels of any (new) participant. Thus, we applied a leave-one-subject-out (LOSO) cross-validation (CV) procedure [12, 8, 51], where a model is trained on the data of all but one participant and evaluated on the data of the remaining participant. Doing so, the average performance of a model represents an estimate of the model’s generalisability to new users.

Here, the MK models were trained on 3574 observations from 36 participants, while the MKH and H models were trained on the 3225 and 2097 valid observations from 32 participants, respectively. Guided by previous literature, we used zero-fill imputation to replace missing values [107, 12], and robust standardisation [8, 84] to scale all features (except the binary sociodemographic features, i.e., work location, gender, and position within the company, which are already coded as 0s and 1s). Note that these preprocessing steps were fitted only on the training data set and then applied to the testing data set for each fold to avoid data leakage.

We considered three different ML algorithms: Elastic Net (EN), Random Forest (RF) and XGBoost (a popular gradient boosting method, [20]). We selected these algorithms due to their successful application in similar research and their ability to handle large feature sets [12, 69, 115, 4]. We tuned the hyperparameters of each algorithm with a grid search (see Appendix C for the details on the chosen grids) while performing the LOSO strategy.

Similar to [12, 101, 39], we used the Spearman rank correlation coefficient, i.e., Spearman’s *ρ*, to evaluate the performance of each ML model during the LOSO CV, and to select to optimal hyperparameter configuration. Spearman’s *ρ* has been proposed as a suitable performance metric for regression models due to its “evaluation robustness” (cf. [98]). In addition, following [121, 99, 68], we also report the mean absolute error (MAE) of the tuned models alongside Spearman’s *ρ* values. Similar to [35, 12], to assess whether a model performed (on average) better than chance, we computed the distribution of the model’s chance performance based on 1000 random shuffles of the stress labels. We infer that a given model shows performance that is significantly higher than chance if it lies above the 95% quantile of this distribution [86]. For an additional baseline comparison, we report the MAE that a dummy model which always predicts the mean value achieves under the implemented LOSO CV procedure. Note that Spearman’s *ρ* cannot be calculated for such a dummy model since the correlation with a constant model is undefined.

##### Time-sequenced features

In the above approach, single, aggregated feature values were derived from the entire 60-minute windows. Here, we used sequences of features on smaller time segments instead. Specifically, we split the 60-minute time window before each EMA report into non-overlapping segments of 5 minutes, similar to [130]. Then, we derived sequences of the same set of mouse, keyboard, and HRV features (described in Sections 4.3.1-4.3.3) on these segments. We considered two sequence lengths: the entire 60-minute window and the last 30 minutes before the EMA report (i.e., sequences of 12 and 6 timesteps, respectively). We then used these sets of features in a one-fits-all modelling approach, i.e., following the LOSO CV procedure. We did not explore the sequenced features for individual models due to the small number of observations available per participant.

Deriving the HRV features on these more granular segments resulted in some additional observations that could be used as input for the H models, namely those which had valid feature values for at least some of the 5-minute segments. However, for other observations, the feature values for the segments in the last 30 minutes were all missing, and these observations were removed based on our logic of removing observations with no predictive information. As a result, for the feature sequences derived on the 60-minute time windows, the MK models were trained on 3574 observations from 36 participants, while the MKH and H models were developed on 3225 and 2668 observations from 32 participants, respectively. Similarly, for the feature sequences derived on the 30-minute time windows, the MK models were trained on 3553 observations from 36 participants, while the MKH and H models were developed on 3208 and 2538 observations from 32 participants, respectively. Following [35, 12, 107], missing values were imputed with zeros after forward-filling within each sequence, and all features were standardised.

Here, we employed Long-Short-Term-Memory networks (LSTM; [52]), a popular type of RNN suitable for sequential data [35, 12]. For comparison with standard methods, we also trained XGBoost models on the same extended set of features. As XGBoost models do not handle sequential data, we trained them on a ‘flattened’ feature vector by concatenating the time-relative features. We also tuned the hyperparameters of these algorithms with a grid search within the LOSO CV procedure (see Appendix C for the grids). Due to the very high dimensionality of features and computational limitations, we did not employ EN or RF methods here. For performance evaluation and baseline comparison, we followed the approach described in 4.4.1.

#### 4.4.2 Personalised approach

In the personalised approach, ML models were trained separately for each participant. To evaluate the individual ML models, we followed [127] and used a blocked CV procedure suitable for longitudinal data. In fact, a standard k-fold CV procedure would violate the independence of train and test splits: two observations that are very close together in time (e.g., from the same day) are arguably more similar than two observations weeks apart, yet, in a standard k-fold CV, the similar observations may likely end up in separate folds [48]. However, in blocked CV, each fold is a temporally continuous block of the data, ensuring that test data stems from a different block of time than the training data. We define a block to be two consecutive weeks of data, which yields four distinct folds based on our study length of eight weeks. The blocks are always separated by two days of the weekend, in line with an *hv*-block CV procedure omitting boundary instances for more rigorous separation [94]. Consequently, we trained each individual model on data from three blocks and evaluated its performance on the fourth block, and repeated the procedure to get an average test performance per participant.

The number of available observations in each block varies over time and per participant. Specifically, we have on average 99.28 observations per participant (SD = 26.77), and 24.82 observations in the respective individual blocks (SD = 8.47). In the personal approach, we used median imputation to replace missing values [11], and robust standardisation [8, 84] to scale all features (except work location), applied within the blocked CV procedure. Note that gender and position within the company are constant for each participant and were not used as features of the ML models.

As in the one-fits-all approach, we considered EN, RF, and XGBoost algorithms. However, because the number of observations was small in relation to the number of features for the individual ML models, we further included a feature elimination step in the modelling pipeline [35]. As in [131, 115], we used recursive feature elimination (with an RF estimator) to iteratively reduce the number of features to 15 before the application of the ML models. Moreover, we explored slightly smaller hyperparameter grids for tuning the individual models to avoid overfitting (see Appendix C).

Analogously to the one-fits-all approach, we tuned and evaluated all models using the Spearman’s *ρ*, and also reported the corresponding MAEs. Since the personalised approach produces a different models for each participant, we report the mean and standard deviation of the Spearman’s *ρ* and MAE across participants. Furthermore, to benchmark against chance, we also compared model performance to the distribution of model performances for shuffled labels with 1000 iterations [35, 86]. Note that here, the labels were shuffled within-participant only. For each algorithm and feature set combination, we counted the number of participants for which the individual models significantly outperformed the chance distributions at an *α* level of 0.05. In addition, we report the MAE that individual dummy models (predicting the mean value for each participant) achieve under the blocked CV procedure, averaged across all participants. Again, Spearman’s *ρ* cannot be calculated for these dummy models since the correlation with a constant model is undefined. Finally, we considered no approach based on time-sequenced features for the individual models, due to the limited number of data points per participant.

## 5 Results

### 5.1 Participant characteristics and user experience

The characteristics of the study participants are summarised in Table 1. As for average levels of chronic stress, a two-sided dependent samples t-test revealed that employees’ self-reported levels significantly decreased from the beginning (Pre: mean = 8.03, SD = 3.72) to the end of the study (Post: mean = 6.61, SD = 4.48), *t*(35) = *−*3.50, *p* = .001. Refer to Table A.1 in the Appendix for an overview of the counts and percentages of the categories of chronic stress levels at Pre and Post.

**Table 1:** Sample characteristics (*N* = 36)

| Variable | $n$ | % |
| --- | --- | --- |
| Sex |  |  |
| Female | 18 | 50.00 |
| Male | 18 | 50.00 |
| Age [mean (SD), range] | 36.67 (11.50) | [18,60] |
| Education |  |  |
| Vocational education | 3 | 8.33 |
| Baccalaureate | 4 | 11.11 |
| College of higher education | 8 | 22.22 |
| University degree | 21 | 58.33 |
| Position |  |  |
| Manager | 11 | 30.56 |
| Employee | 25 | 69.44 |
| Workload |  |  |
| 100% | 19 | 52.78 |
| 90% | 5 | 13.89 |
| 80% | 7 | 19.44 |
| 70% | 2 | 5.56 |
| 60% | 3 | 8.33 |

Abbreviations: SD = Standard deviation.

Overall, most participants (31/36) rated the system usability of the data collection app as good to excellent (SUS score: mean = 81.00, SD= 12.13 on a scale from 0 to 100). In terms of obtrusiveness, 9 out of 36 participants rated wearing the armband daily as obtrusive in their daily life, while 11 out of 36 participants felt the EMA prompts were obtrusive. All participants found the mouse and keyboard data collection to be unobtrusive.

### 5.2 Self-reported stress

In total, participants were prompted 4585 times by the software to report their psychological state and provided 3574 completed responses, corresponding to a response rate of 77.95%. On average, we received 99.28 self-reports per participant (SD = 26.77, range = [49, 150]). Participants answered between 1 and 8 EMAs per day (mean = 3.32, SD = 1.02), and had between 17 to 41 distinct data collection days (mean = 29.89, SD = 5.85). The average stress score was 27.15 (SD = 23.44). See Figure 2 for the distribution of the recorded self-reports values.

**Figure 2:**
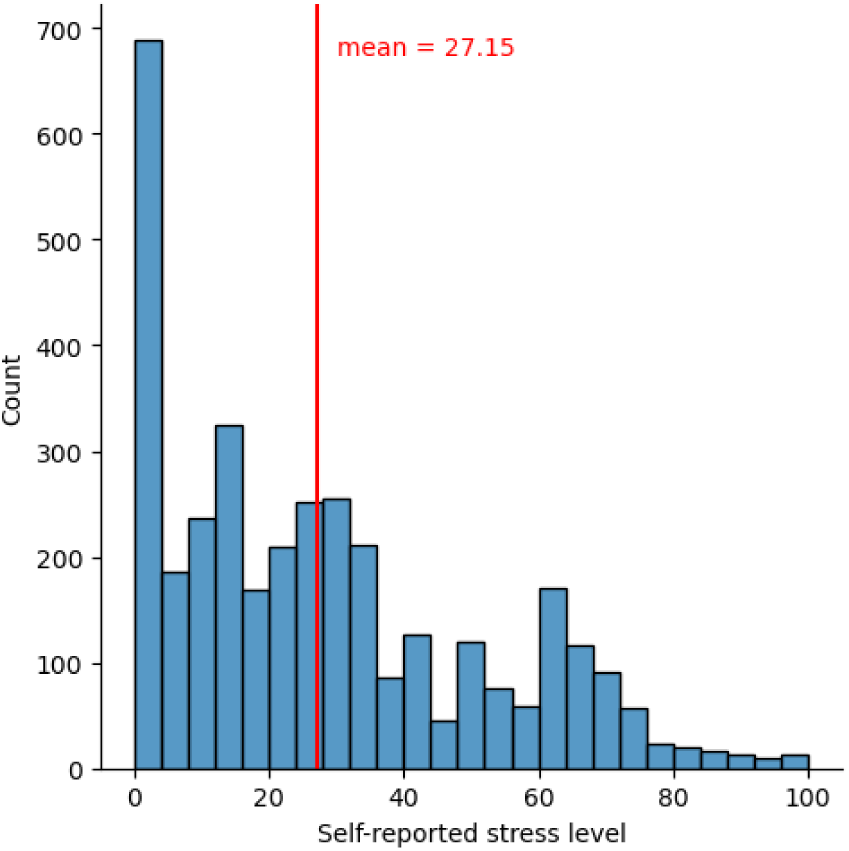
Histogram of the stress levels recorded via Ecological Momentary Assessments. Higher values on the *x*-axis indicate higher levels of self-assessed stress.

### 5.3 Machine learning modelling results

#### 5.3.1 One-fits-all approach

Table 2 provides an overview of the best-performing EN, RF, and XGBoost models for the different feature subsets under the one-fits-all approach. Table 3 contains the results for the LSTM and XGBoost models on the time sequences of features.

**Table 2:** Performance results of models following the one-fits-all approach, evaluated using leave-one-subject-out cross-validation.

| | | Spearman’s $\rho$ | | | MAE | |
| --- | --- | --- | --- | --- | --- | --- |
| | | mean | (SD) | $p$-value | mean | (SD) |
| Elastic Net | MKH | 0.039 | (0.123) | .052 | 19.43 | (6.12) |
|  | MK | 0.045 | (0.125) | .019 | 19.58 | (5.78) |
|  | H | 0.038 | (0.160) | .100 | 19.58 | (6.25) |
| Random Forest | MKH | 0.056 | (0.107) | .007 | 20.68 | (6.23) |
|  | MK | 0.042 | (0.117) | .017 | 20.81 | (6.54) |
|  | H | 0.030 | (0.139) | .175 | 19.75 | (6.14) |
| XGBoost | **MKH** | **0.078** | **(0.155)** | **<.001** | **19.99** | **(6.33)** |
|  | MK | 0.051 | (0.140) | .012 | 20.75 | (6.35) |
|  | H | 0.063 | (0.143) | .032 | 19.34 | (8.98) |
| Mean baseline | MKH |  | - |  | 19.19 | (6.10) |
|  | MK |  | - |  | 19.64 | (5.84) |
|  | H |  | - |  | 19.49 | (6.26) |

Abbreviations: Spearman’s $\rho$ = Spearman’s rank correlation coefficient; MAE = mean absolute error. SD = Standard deviation; $p$ -values represent the percentage of the cases in which the mean Spearman’s $\rho$ of a given approach outperformed the corresponding randomly shuffled baseline estimators; M = mouse features; K = keyboard features; H = heart rate variability features.

**Table 3:**
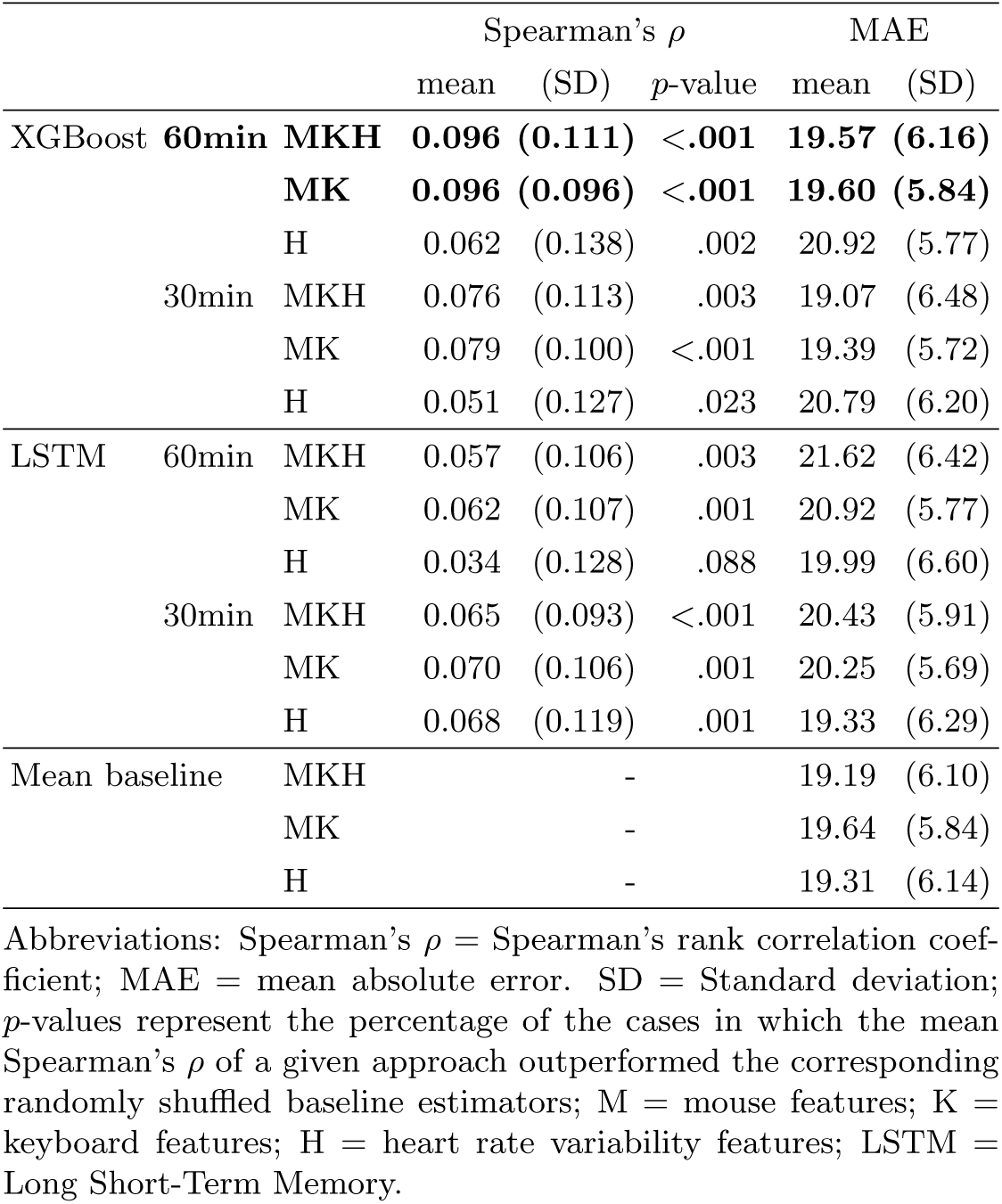
Performance results of models following the one-fits-all approach and trained on time-sequenced features, evaluated using leave-one-subject-out cross-validation.

Overall, the results reveal weak correlations of the predicted values with the ground truth self-reported stress levels across algorithms, feature sets and approaches. In the standard one-fits-all approach, the highest mean Spearman’ *ρ* is achieved by the XGBoost model based on mouse, keyboard, and HRV feature data at 0.078. Using time sequenced input features rather than the one-hour aggregations yields small improvements. In particular, the best performance is reached by the XGBoost algorithm trained on the feature sequences derived on 60 minute windows. Here, the XGBoost models based on mouse, keyboard data with or without the addition HRV data both reach the highest Spearman’ *ρ* of 0.096, corresponding to a 23% increase compared to the performance of the standard one-fits-all approach.

In terms of algorithms, XGBoost models reach the highest performance levels, while the RF and EN models reach similar results. XGBoost also outperforms the LSTM models for the time sequenced features, especially on the longer 60 minute time windows. Regarding the different feature sets, the full array of features works best in the case of RF and XGBoost models at the one hour aggregation level, but for the time sequence features, the models using on only mouse and keyboard data yield performances just as high or slightly better than the MKH models. By contrast, the cardiac data models generally perform worse than both MKH and MK models.

The standard deviations in performance results are also consistently high, implying that models on some test folds (i.e., subjects) reached substantially higher but also lower performance than the reported averages. In addition, even the best-performing models in terms of achieved Spearman’s *ρ* exhibit similar or slightly higher MAE of predictions compared to a dummy model predicting the mean. Notably, the *p*-values of the model performances compared against the distributions from the respective null models trained from shuffled labels are below 0.05 in most cases, indicating that these models managed to learn at least significantly more than spurious effects.

#### 5.3.2 Personalised approach

We report the results of the personalised approach in two ways: (1) by ML model family, aggregated across participants, and (2) by selecting the best-performing ML model individually for each participant.

##### Performance of the personalised approach by ML model family across participants

Table 4 presents the performance results of individual models trained using a blocked CV procedure, aggregated across participants, for each ML model family. The highest Spearman’s *ρ* averaged across participants is achieved by the XGBoost model family trained on mouse and keyboard features at 0.188. Here, the MK models slightly outperform the other feature sets for all algorithms, while XGBoost again obtains somewhat higher performance than EN and RF models. The standard deviations of performances across participants are also high, suggesting rather high inter-participant performance variability. The MAEs of the personalised models also show no improvement with respect to the MAEs of the dummy models predicting the mean stress level for each participant. Finally, the comparisons with the performances achieved by null models on shuffled data show that for the best-performing XGBoost MK models, at least for more than half of participants the individual models achieved higher scores than 95% of chance-level models.

**Table 4:** Performance of the personalised approach by ML model family across participants. Individual models are evaluated using blocked cross-validation, reported values are averaged across participants.

| | | Spearman’s $\rho$ | | | MAE | |
| --- | --- | --- | --- | --- | --- | --- |
|  |  | mean | (SD) | N sign. | mean | (SD) |
| Elastic Net | MKH | 0.124 | (0.127) | 8/32 | 13.73 | (7.19) |
|  | MK | 0.155 | (0.126) | 13/36 | 14.52 | (7.30) |
|  | H | 0.102 | (0.156) | 4/32 | 13.77 | (7.50) |
| Random Forest | MKH | 0.125 | (0.148) | 9/32 | 13.58 | (7.00) |
|  | MK | 0.148 | (0.151) | 10/36 | 14.07 | (6.73) |
|  | H | 0.105 | (0.151) | 3/32 | 14.07 | (7.45) |
| XGBoost | MKH | 0.178 | (0.129) | 13/32 | 14.51 | (7.38) |
|  | **MK** | **0.188** | **(0.122)** | **19/36** | **15.17** | **(7.50)** |
|  | H | 0.185 | (0.115) | 6/32 | 15.05 | (7.45) |
| Mean baseline | MKH |  | - |  | 13.07 | (7.28) |
|  | MK |  | - |  | 13.81 | (7.27) |
|  | H |  | - |  | 13.21 | (7.45) |

Abbreviations: Spearman’s $\rho$ = Spearman’s rank correlation coefficient; MAE = mean absolute error. SD = Standard deviation; N sign. represent the number of participants for which the mean Spearman’s $\rho$ of a given approach outperformed 95% the corresponding individual randomly shuffled baseline estimators; M = mouse features; K = keyboard features; H = heart rate variability features.

Further, we investigated the most important features of the XGBoost model on mouse and keyboard data. Specifically, we counted which features appeared most frequently in the list of top-10 most important features for the individual models, similarly to [39], see Table 5. Results reveal that no feature seems to be highly important across the majority of the (best) ML models. Rather, the trigraph-related features were among the most important for around one-fourth of the participants, while mouse movement features relating to the traversed angle and the cursor acceleration were also selected for roughly one-fifth of participants.

**Table 5:** Features that most frequently rank among the top 10 in the per-participant best XGBoost models on mouse and keyboard data (see. **Table 4).**

| Feature name | Description | Count |
| --- | --- | --- |
| ICH_Dur2Mean | Mean of the duration of second key in the ‘ICH’ trigraph | 9 |
| NDE_Btw23Mean | Mean of the duration between second and third key in the ‘NDE’ trigraph | 8 |
| AvgSignAngleMean | Mean of the average (unsigned) angles traversed within a mouse movement | 7 |
| ICH_Dur1Mean | Mean of the duration of first key in the ‘ICH’ trigraph | 7 |
| ICH_Dur3Std | standard deviation of the duration of third key in the ‘ICH’ trigraph | 7 |
| ScrollDownCount | Number of scrolling down operations | 7 |
| StdAccelNegStd | standard deviation of the variation in negative acceleration within a mouse movement | 7 |
| StdAccelPosStd | standard deviation of the variation in positive acceleration within a mouse movement | 7 |

Abbreviations: M = mouse features; K = keyboard features; Count represents the number of participants for which the corresponding feature was among the top 10 most important features.

Finally, we studied the correlation between the performance of the per-participant best XGBoost models on mouse and keyboard data and the socio-demographic characteristics of each participant. As is evident in Figure 3, the achieved Spearman’s *ρ* scores of the individual models vary greatly across participants, with two participants reaching negative values. In Figure 4, we plot a number of participant characteristics against the performance of these models. However, the plots reveal no evident trends.

**Figure 3:**
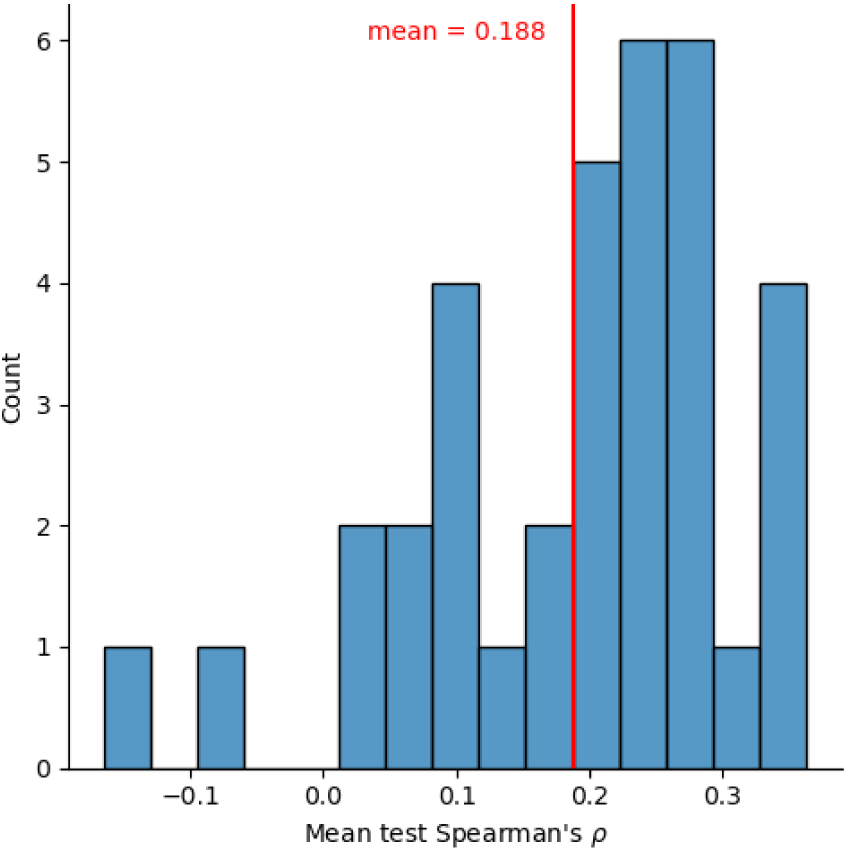
Personalised approach: Distribution of the mean test performance of the best per-participant XGBoost models on mouse and keyboard data–see Table 4.

**Figure 4:**
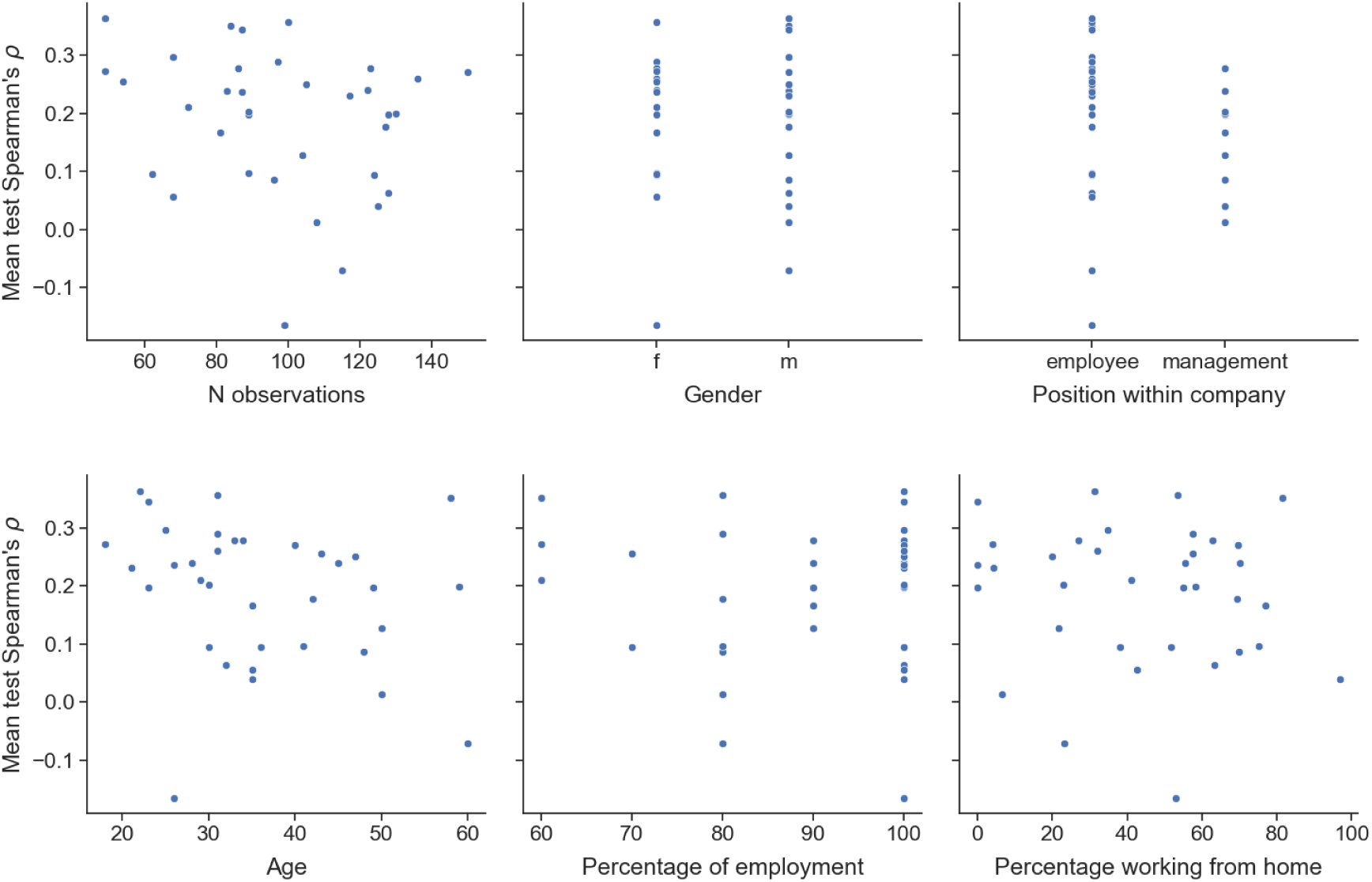
Personalised approach: Mean test performance of individual models for the XGBoost mouse and keyboard-based models plotted against different participant characteristics.

##### Performance of the personalised approach by best ML model per each participant

We close the analysis of the personalised-approach by collecting the best ML models per participant (note that different participants may have different best models) and investigating the distribution of their performance. We show the distribution and the best models–including their data modalities–in Figure 5. The mean Spearman’s *ρ* equals 0.296 (SD = 0.083), corresponding to a 57% increase compared to the average performance of the personalised approach by ML model family across participants. Top performance is achieved by an RF model on mouse and keyboard data and it is equal to *ρ* = 0.462. In total, 16 our of 36 models (44%) reach performance above the mean *ρ* = 0.296. Finally, six out of 36 models (17%) reach performance above *ρ* = 0.400; they are two RF models (both on mouse and keyboard data), followed by two XGBoost models on cardiac data only, followed by a XGBoost model on mouse, keyboard, and cardiac data and an RF model on mouse and keyboard data.

**Figure 5:**
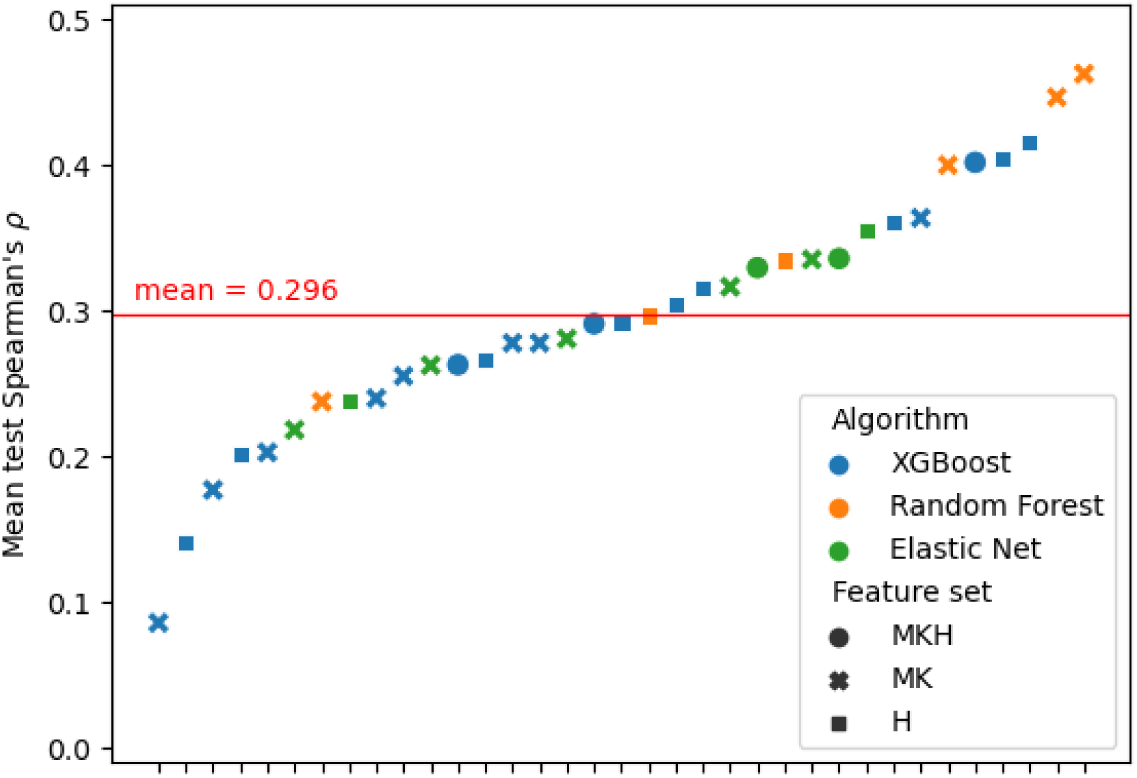
Personalised approach: Distribution of the best model performance per participant. The best models change across participants. Abbreviations: M = mouse, K = keyboard, H = heart rate variability.

## 6 Discussion

In this work, we present the results of an 8-week field study in which we collected data on employees’ mouse and keyboard usage and cardiac activity during working hours. To the best of our knowledge, this is the first longitudinal field study to investigate the combination of mouse movement characteristics, keystroke dynamics and HRV features for stress detection. The collected data were used to train ML regression models predicting employees’ psychological state, which they reported several times a day. The data set is available online in the OSF repository at https://osf.io/qpekf/; access will be granted upon request.

The study sample consists of 36 employees aged between 18 and 60, and fairly balanced in terms of gender, position within the company, and percentage of employment. Participants also spent varying percentages of their time working remotely. The comparison of chronic stress levels assessed by the TICS revealed that participants generally had lower levels of chronic work-related stress after the data collection than before. Since this was not an intervention study and as such did in particular not involve any sort of control group, we cannot make any claims as to whether this effect could be attributed to the data collection itself. While the numbers indicate that using the system did not further contribute to employees’ stress levels, some research has found that monitoring one’s mental health can lead to improvements in self-awareness and symptoms [81, 58].

The distribution of the daily, self-reported stress labels is slightly bimodal, shows a heavy skew towards the left and considerable variance both between and within participants. As Booth et al. [12] argue, quantising this scale into binary or ordinal labels by some arbitrary threshold could artificially eliminate this variance, and reduce the convergent validity of derived models. We therefore followed a regression approach to detect participants’ self-reported stress levels from mouse, keyboard and cardiac data.

### 6.1 One-fits-all approach

We found that one-fits-all modelling approaches reached low performances in combination with a LOSO CV procedure. While the highest obtained Spearman’s rank correlations surpass chance level, the achieved values are in the negligible to low range in terms of effect size, and the MAEs of the best models remained below those of a constant mean estimator.

Furthermore, while it is difficult to directly compare performance results between stress detection field studies due to differences in study design, collected data, derived features, ML methods, performance measures, etc., we try to contextualise our results with a selection of similar works. Likely the most comparable work in terms of data modalities, in [82] the authors collected mouse, keyboard, and HRV data (among other physiological and contextual data sources). However, their mouse and keyboard data is limited to the total number of mouse clicks, wheel rotations and pressed keys and does not include any mouse movement or keystroke dynamics-related features. The authors report an F1-score of 46% under LOSO CV when detecting low versus high perceived stress. Soto et al. [115] also aimed to detect two classes of stress based on a field study collecting employees’ HRV and other physiological data with a wearable. Their one-fits-all modelling approach is outperformed by both a majority and a random baseline classifier. Similar to our own findings, both studies thus achieved low performance values following one-fits-all approaches for stress detection in the field—even while considering only binary classification. In contrast, Booth et al. [12] also followed a regression modelling approach. With their meticulous effort to develop models which safeguard both robustness and validity, the authors achieved Spearman’s rank correlations of up to 0.18 with self-reported daily stress levels under nested LOSO CV, based on HRV measures as well as sleep, activity, location, phone usage and weather data collected from 606 employees. Interestingly, while Booth and colleagues [12] found that time-aware methods such as LSTM and Gated Recurrent Unit (GRU) could not reach better than chance performance, our results indicate that RNN may be able to offer small performance improvements. Nevertheless, the XGBoost models on time-sequenced features outperformed the LSTM models in our study. While the limited number of available data points could be favouring the boosting-based approach over more data-intensive neural nets, this result is in line with the observed trend according to which tree-based methods are superior in performance with respect to deep learning ones on tabular data [47].

### 6.2 Personalised approach

The personalised approach yielded generally higher performance levels than the one-fits-all models when considering the best ML model families across participants. Still, the obtained Spearman’s rank correlations can be considered small, and we observe that for the mouse and keyboard-based XGBoost models, which achieve the highest Spearman’s *ρ* on average, slightly more than half of the individual models performed better than chance. Moreover, their average MAE is higher than the one of individual mean estimator models. However, when selecting the best ML model for each participant out of all feature sets and algorithms in the personalised approach, we achieved an average Spearman’s *ρ* of 0.296, reflecting a close to medium effect size and surpassing the performance of Booth et al.’s (2022) one-fits-all approach [12]. Forty-four percent of the best models achieve Spearman correlations within the medium effect size range, and 17% exceed a correlation of 0.400. Further, the highest individual score even rises up to 0.462 for one participant reflecting a medium-to-large effect.

These results provide encouraging evidence for the viability of a personalised modelling strategy that focuses on the most suitable ML model and data modality for each individual. As highlighted by Booth et al. [12], small to medium-sized effects—such as those observed in our study—are consistent with longstanding findings in psychological research and typical of associations between behavioural signals and subjective mental states, especially in naturalistic settings [28]. At the same time, the variability across participants in both optimal algorithms and data modalities—e.g., some participants benefitting more from mouse and keyboard features, others from HRV—indicates the need for further investigation into individual differences and refinement of the experimental design to better account for such heterogeneity.

While, to our knowledge, no stress detection field studies exist which follow a personalised approach and involving mouse, keyboard and cardiac data, the previously mentioned work by Soto et al. [115] reports an average accuracy of 0.768 with individual models based on HRV and EDA data. Representing only a small improvement over the 0.735 average accuracy of majority class estimators, the individual models obtained at least moderate improvements (25.9%) in precision for the “stressed” class.

Our analyses further revealed that socio-demographic characteristics did not correlate with the individual model performances. In contrast, Smets et al. [112] found that participants with higher performances had on average more imbalanced stress labels, a higher number of provided feedbacks, and a higher dynamic range in physiological feature values. This last finding might imply that some people have more discernible physiological responses to stress. Similarly, our investigation of the best models per individual (out of all algorithms and feature combinations) revealed that HRV-based models achieved the highest scores for some participants. Here, psychophysiological research suggests that underlying individual psychological traits and states and physiological dispositions can affect the degree of correspondence between physiological and psychological stress measures [14].

### 6.3 Data modalities

The low performance results for the mouse and keyboard-based models in the one-fits-all approaches support the findings from previous laboratory experiments, which found no generalisable association between mouse or keyboard usage and self-reported stress [35, 36]. However, in line with [128], we found that personalised approaches show potential to detect stress levels. Furthermore, we found that the models based on HRV data usually performed worse than the mouse and keyboard-based models in either approach, despite the theoretically and empirically well-established link between HRV and stress [9, 90, 61]. One possible interpretation is that the data quality of the collected PPG data in our study was simply too low, leading to too many noisy or invalid feature values. Here, Menghini et al. [78] note that keyboard typing can lead to a significant amount of artefacts in PPG-based measures. Indeed, in our study, we observed that participants had missing HRV features data for 35.36% of their observations on average (SD = 14.04%, range [11.90%, 62.61%]). Another explanation could be that in the field, under the influence of confounders such as physical movements and cognitive load, the link between HRV and psychological stress is not specific enough to be detected easily. This interpretation would be in line with previous findings from a lab study simulating an office environment where participants performed baseline workload tasks and stress was elicited by work-related stressors [84]. There, the authors found that models based on mouse and keyboard data outperformed models based on cardiac data. It has also been shown that there is often a dissociation between the psychological and the physiological stress response due to underlying moderating or mediating factors or inter-individual differences [14]. Notably, recent stress detection studies collecting HRV data in the field also report low performances [12, 82, 115], and a recent investigation by Martinez et al. [74] found HRV to be only weakly associated with perceived stress under real-world conditions.

### 6.4 Study Limitations and Recommendations for Future Work

In the following, we provide a structured discussion of the main limitations that affect research on the automated detection of stress with machine learning in real-world settings, and we give actionable recommendations for future investigations in that field.

#### Reliable collection of cardiac, mouse, and keyboard data in real-world field settings

As discussed in Section 6.3, PPG data collected in real-world settings—where employees are engaged in activities such as moving, typing, talking, and performing mentally demanding tasks—appear to face significant quality issues when using current consumer-grade wearables, as observed in our study and the literature [78, 30]. In particular, ambulatory HRV measurements are susceptible to motion artifacts and environmental conditions (e.g., temperature, moisture) that may degrade signal quality [111]. Thus, better sensor technology and improved signal processing techniques to handle motion artefacts as well as noise in the extraction of inter-beat intervals from raw data are needed. Further, mouse movement data, as demonstrated in our study, can be collected more reliably but often in large volumes, requiring streamlined preprocessing pipelines to efficiently extract movement characteristics. These pipelines must accommodate variations in screen setups and account for transitions between different input devices, such as mice and trackpads. Finally, our collection of keyboard data was somewhat limited by the local aggregation of raw data and minor errors in the data collection software (cf. Section 4.3.2). Here, more fine-grained and improved aggregation protocols could potentially lead to better quality raw data, while still mitigating potential security and privacy concerns of employees and employers [59].

#### Missing data

While many knowledge workers spend the majority of their day interacting with a computer, there are inevitably periods when only a mouse or keyboard is used, or when no activity from these devices is recorded. Additionally, PPG devices may experience functionality issues leading to missing data, as evidenced in our study, where participants had an average of 35.36% missing HRV feature data across their observations. Therefore, strategies to handle missing data are crucial. Possibilities range from simple constant imputation schemes, including zero-fill, mean-fill, person-specific mean-fill [12], to more sophisticated approaches. For instance, Jaques et al. [55] developed a specialised multimodal autoencoder for handling missing data and enabling more robust mood prediction. However, more research is needed to determine which approaches are most beneficial in the context of stress detection based on physiological and behavioural data sources.

#### Stress label assignment

Stress detection field studies vary considerably in the duration of time windows used for feature calculation and their alignment with EMA-based stress labels. In our study, we selected a one-hour time window to align with the phrasing of the EMA question, “How stressed did you feel in the last hour?” following EMA guidelines [109]. Other studies have used time windows ranging from seconds or minutes [115, 17] to several hours or even entire days [101, 12]. Additionally, while most studies define time windows to immediately precede EMA reports, Gjoreski et al. [44] also explored extending the window up to 10 minutes after the EMA entry, assuming that the reported psychological state might persist during this period. Future research should carefully evaluate the choice of temporal resolution for stress labels, considering both psychological theory and the intended practical applications of the developed models.

#### Ecological validity of data collection at the workplace

A key prerequisite for advancing research in automated stress detection in real-world settings is the collection of large, ecologically valid datasets. Our study comprised N=36 participants from a single company and a duration of eight weeks. This resulted in our data set of 3574 labelled observations, which remains a relatively small amount for the application of machine and, in particular, deep learning methods. Here, the importance is not just on greater and more diverse subject pools (e.g., cohorts from different companies and sectors) but also on acquiring data over longer periods of time per participant. Especially for individual models, the study length should be long enough to capture the entire range of a person’s ecologies, including seasonalities, vacation periods, and other events, such as illness or personal leaves [95]. Further, in our study, we did not collect information regarding participants’ workstation setups (e.g., the number and positioning of screens they used, whether they employed the laptop keyboard and trackpad or external devices) and whether they might alternate between different setups (e.g., at home versus in the office). To this end, to improve ecological validity of data collection, future research needs to incorporate additional setting-specific data, such as background noise levels, traffic and weather conditions, work location, task type, and work-related stressors like email overload, unplanned meetings, or distractions [12]. Moreover, the collection of ground truth labels represents one of the main limiting factors. While EMAs offer a powerful tool to collect ecologically valid data at scale, their design needs to be concise and brief enough to limit the burden on participants and assure compliance with collection [96]. Furthermore, while researchers may wish to trigger EMA prompts as frequently as possible to capture diverse experiences and contexts, too many prompts or prompts at inopportune moments may lead to response fatigue and inaccurate or missing self-reports [42]. Here, Ghosh et al. [42] designed a schedule for smartphone-based EMA collection aimed at reducing prompt rate, improving timely self-report collection and eliminating inopportune prompts. Adapting such an approach to work environments could be particularly beneficial to minimise interrupting employees in their workflows or in critical moments (e.g., during meetings). Therefore, future research should investigate the most reliable and efficient EMA designs for stress assessment, both in terms of questionnaire content and prompt schedule. Alternatively, researchers could address the scarcity of labelled data through ensemble methods, semi-supervised learning, and transfer learning, as proposed by Maxhuni et al. [76]. In addition, more field studies should strive to make their collected data sets public, such as TILES [83], StudentLife [126], and the one presented in this work.

#### More robust machine learning personalised approaches

Our results indicate that personalised models may be a more promising avenue for future research compared to one-fits-all approaches. Developing and managing individual ML pipelines for all subjects (e.g., a cohort of employees in a corporate work environment) requires appropriate resources and necessitates the collection of sufficient training data for any new user before the stress detection system can be deployed. Therefore, the use of personalised approaches for the automated detection of stress at work depends on the scalability of the resources required to manage all models and the users’ willingness to undergo an extensive data collection procedure before being able to reap the benefits stemming from the automated detection of their stress levels. A number of studies have explored alternatives which compromise between individual versus general models. Examples are the development of models for clusters of similar users [16], a “mixed” modelling approach where individual models are augmented with training data from other users [127], the use of more data-intensive deep learning approaches such as transfer learning [76] or multitask learning [117]. While the results from these work generally show promise for the different approaches, they have yet to be investigated in the context of mouse and keyboard-based stress detection models. However, the performance of personalised models, including those developed in this study, is not yet sufficient for practical application in real-world settings. Future research should further investigate the use of contextual information to enhance modelling performance in automated stress detection. For example, Booth et al. [12] demonstrated that including participants’ stress-related states, such as anxiety measured through the State-Trait Anxiety Inventory questionnaire, significantly improved the performance of their one-fits-all approach. Additional contextual data, such as participants’ socio-demographic characteristics (e.g., family structure) beyond basic details like age, gender, and employee status, as used in our study, could further improve the models. Contextual information not only enhances ecological validity but has also been shown to improve performance in automated stress detection systems [79, 44]. Finally, while we employed LOSO CV and blocked CV procedures to obtain estimates for how our models would perform on data from other participants and other weeks, respectively, nested CV procedures could provide even more robust and unbiased assessments of performance and generalisability.

#### Hand-crafted features versus deep learning methods

The approach followed in this work and in the majority of similar studies relies on hand-crafting features from raw data [105]. While the HRV measures are grounded in psychophysiological research and established markers of ANS reactivity to stress [9], the engineering of keystroke dynamics and mouse movement features has no such theoretical foundation. Consequently, alternative representations of mouse and keyboard data may be able to detect stress levels with better performance. For example, Freihaut [35] exploratively investigated using visualisations of mouse trajectories as input for a Convolutional Neural Net (CNN) and using raw cursor coordinates time series as input for an LSTM, although neither approach yielded satisfactory performance. In a follow-up study, Freihaut et al. [37] used the CNN Resnet 34 on images of mouse trajectories and clicks obtaining similar results. However, more recently, Quadrini et al. [93] introduced a method that encodes physiological signals into images via Markov transition fields for the classification of stress using CNNs and tested it on multiple datasets, outperforming alternative approaches. In addition, deep learning methods applied to raw mouse movement and keystroke data have in recent years gained much attention in user authentication research [21, 92], the field which has also served as inspiration for many of the existing mouse and keyboard feature definitions. Regarding cardiac data, some researchers also achieved improved stress detection accuracies with deep learning models applied raw ECG data under laboratory conditions [105, 31]. However, Pimentel et al. [91] found that simpler ML methods with feature engineering outperformed deep learning methods on HRV and breathing data in the field. These results suggest that more research is needed to investigate the full potential of deep learning methods for stress detection. Importantly, while these methods require substantial amounts of training data, they can easily be combined with transfer or multitask learning paradigms to allow for increased personalisation.

## 7 Conclusion

In this work, we present results from an 8-week field study collecting mouse and keyboard usage and cardiac data from corporate employees at work. Using mouse and keyboard activity and HRV features, we train different regression models to detect self-reported stress levels, both in a one-fits-all and a personalised approach. Our results show that, although one-fits-all approaches show weak correlations with self-reported stress levels and time-sequenced modelling seems to yield limited improvements, the small to medium correlations delivered by personalised modelling approaches offer a more reliable way forward. Similarly to Booth et al. [12], we contend that reliably detecting perceived stress in real-world settings through passive sensing remains an open challenge. This study can lay a foundation for future research exploring the use of multimodal data sources to enhance our understanding of stress in naturalistic environments.

## Data Availability

Data are added to a repository in the Open Science Framework (OSF) under https://osf.io/qpekf/ and are available upon request.

## Acknowledgements

We thank Dr. Erika Meins and Dr. Stefan Wehrli for their support in facilitating this study. We thank the corporate communications, IT and legal departments of the company where the study was conducted for their assistance in the realisation of the study and all the company’s employees who took the time to participate and contribute to our research. At the time of this study, Andrea Ferrario was an employee of the Mobiliar Lab for Analytics at ETH Zurich, whose generous support he gratefully acknowledges.

## CRediT authorship contribution statement

**Mara Naegelin**: Conceptualization, Methodology, Software, Formal analysis, Investigation, Data Curation, Writing – original draft, Writing – review & editing, Visualization, Project administration, Funding acquisition. **Raphael P. Weibel**: Conceptualization, Methodology, Software, Investigation, Data Curation, Writing – review & editing, Project administration, Funding acquisition. **Jasmine I. Kerr**: Conceptualization, Methodology, Investigation, Data Curation, Writing – original draft, Writing – review & editing, Project administration, Funding acquisition. **Florian von Wangenheim**: Resources, Writing – review & editing, Supervision, Funding acquisition. **Victor R. Schinazi**: Writing – review & editing, Funding acquisition. **Roberto La Marca**: Writing – review & editing, Funding acquisition. **Christoph Hoelscher**: Writing – review & editing, Funding acquisition. **Urs M. Nater**: Writing – review & editing. **Andrea Ferrario**: Conceptualization, Methodology, Formal analysis, Writing – review & editing, Supervision, Funding acquisition.

## Appendix

### A Chronic stress levels

See Table A.1 for the overview of reported chronic stress levels among participants.

**Table A.1:** Chronic stress levels [SSCS TICS-2-K].

| Categories | Pre | Post |
| --- | --- | --- |
| Below-average to average | 9 (25.00%) | 18 (50.00%) |
| Above-average | 21 (58.33%) | 12 (33.33%) |
| High | 6 (16.67%) | 6 (16.67%) |

*Note.* Below-average to average chronic stress: 0 to 5 points, above-average levels of chronic stress: 6 to 11 points, high chronic stress: 12 to 24 points. Abbreviations: SSCS TICS2-K = Screening Scale of the short version of the Trier Inventory for the Assessment of Chronic Stress (TICS-2-K, score range: 0 to 24).

### B Feature descriptions

Tables A.2, A.3 and A.4 list and describe the features used in the stress detection models.

**Figure A.1:**
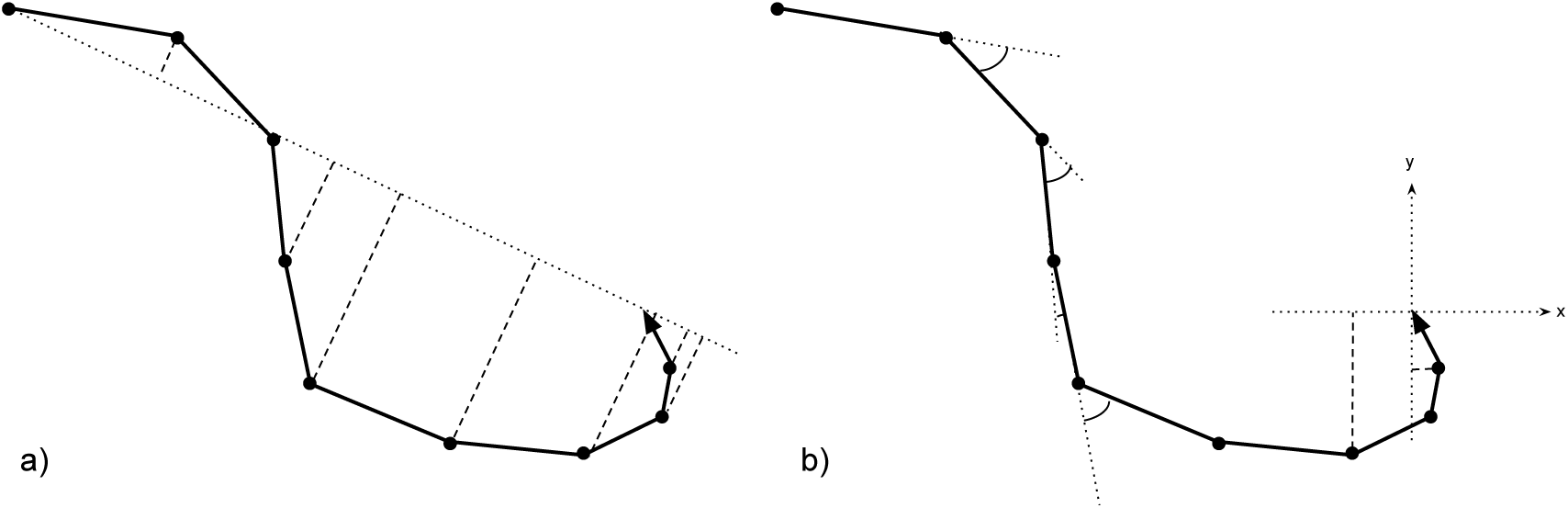
Schematic visualisation of a mouse movement, adapted from [84] with permission. a) The dashed lines indicate the distances from the optimal straight line to the recorded trajectory. b) The arcs indicate the angles traversed along the way, while the dashed lines indicate projections onto the *x* and *y* axes.

### C Hyperparameter grids

See Tables A.5, A.6 and A.7 for the hyperparameter grids of each modelling approach.

**Table A.2:**
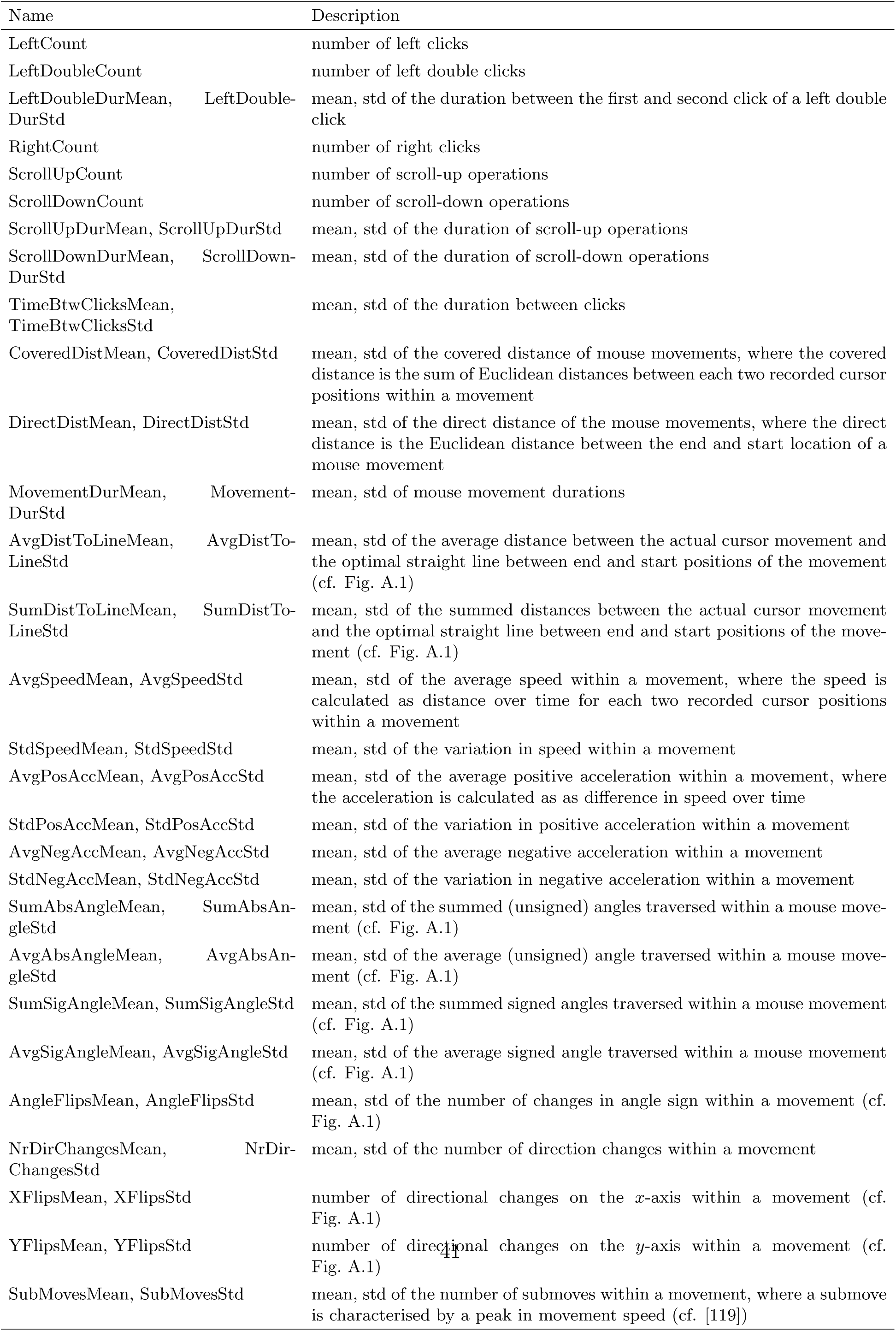
Mouse feature descriptions.

**Table A.3:**
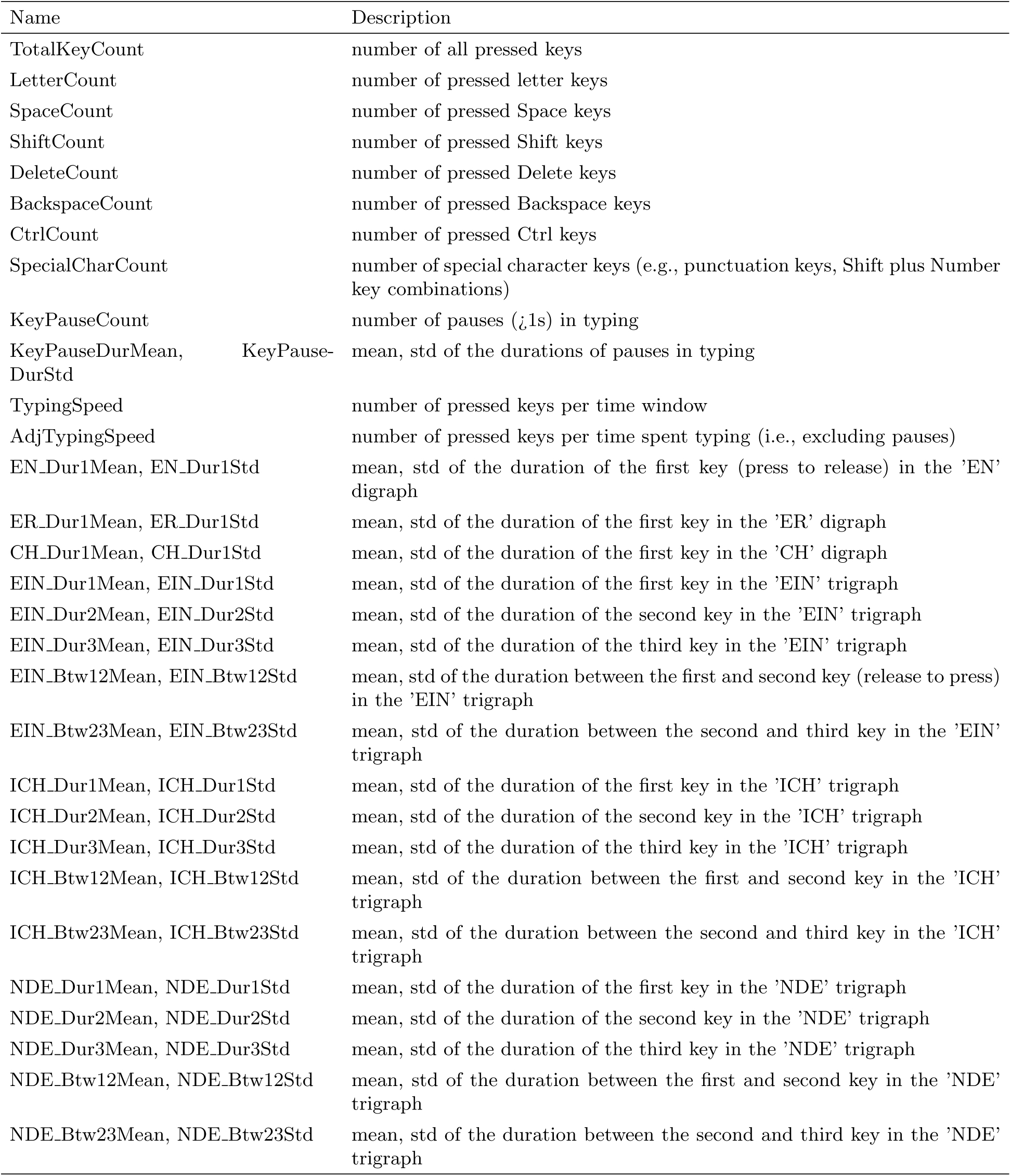
Keyboard feature descriptions.

**Table A.4:** HRV feature descriptions. Abbreviations: IBI = inter-beat-intervals.

| Name | Description |
| --- | --- |
| MeanHR | mean heart rate |
| RMSSD | root mean square of successive differences of IBI |
| SDNN | standard deviation of IBI |
| pNN50 | percentage of successive differences of IBI that differ by more than 50 ms |
| VLF | power in the very low frequency band (0.0033–0.04 Hz) of the power spectral density of the IBI time series |
| LF | power in the low frequency band (0.04–0.15 Hz) of the power spectral density of the IBI time series |
| HF | power in the high frequency band (0.15–0.4 Hz) of the power spectral density of the IBI time series |
| LF/HF | ratio of the power in the low frequency band over the power in the high frequency band of the power spectral density of the IBI time series |
| SD2 | standard deviation along the line of identity of the Poincaré plot of the IBI |

**Table A.5:** One-fits-all approach: Hyperparameter tuning grids for each algorithm.

| Algorithm | Hyperparameter name | Range |
| --- | --- | --- |
| Elastic Net | alpha | 0.1, 0.2, 0.5, 1.0, 2.0, 5.0, 10.0 |
|  | l1_ratio | 0.001, 0.01, 0.1, 0.2, 0.5, 0.7, 0.9 |
| Random Forest | number of trees | 100, 200, ..., 500 |
|  | tree depth | 3, 5, 7, 10, 12 |
|  | maximum number of features (percentage) | 0.6, 0.8, 1.0 |
| XGBoost | number of trees | 100, 200, ..., 500 |
|  | tree depth | 3, 5, 7, 10, 12 |
|  | column subsampling by tree | 0.6, 0.8, 1.0 |
|  | learning rate | 0.01, 0.1, 0.3 |

**Table A.6:** One-fits-all approach, time-sequenced features: Hyperparameter tuning grids for each algorithm.

| Algorithm | Hyperparameter name | Range |
| --- | --- | --- |
| XGBoost | number of trees | 100, 200, ..., 500 |
|  | tree depth | 3, 5, 7, 10, 12 |
|  | column subsampling by tree | 0.4, 0.6, 0.8 |
|  | learning rate | 0.01, 0.1, 0.3 |
| LSTM | number of layers (depth) | 1, 2, 3, 4 |
|  | number of units (width) | 10, 20, 30, 40 |
|  | optimizer | Adam, SGD |
|  | learning rate | 0.001, 0.01, 0.1 |

**Table A.7:** Personalised approach: Hyperparameter tuning grids for each algorithm.

| Algorithm | Hyperparameter name | Range |
| --- | --- | --- |
| Elastic Net | alpha | 0.1, 0.2, 0.5, 1.0, 2.0, 5.0, 10.0 |
|  | l1_ratio | 0.001, 0.01, 0.1, 0.2, 0.5, 0.7, 0.9 |
| Random Forest | number of trees | 50, 100, 150, 200 |
|  | tree depth | 3, 5, 7, 10 |
|  | maximum number of features (fraction) | 0.6, 0.8, 1.0 |
| XGBoost | number of trees | 50, 100, 150, 200 |
|  | tree depth | 3, 5, 7, 10 |
|  | column subsampling by tree | 0.6, 0.8, 1.0 |
|  | learning rate | 0.01, 0.1, 0.3 |

